# A data-driven framework for clinical decision support applied to pneumonia management

**DOI:** 10.1101/2023.07.19.23291197

**Authors:** Robert C. Free, Daniel Lozano Rojas, Matthew Richardson, Julie Skeemer, Leanne Small, Pranabashis Haldar, Gerrit Woltmann

## Abstract

Despite their long history, it can still be difficult to embed clinical decision support into existing health information systems, particularly if they utilise machine learning and artificial intelligence models. Moreover, when such tools are made available to healthcare workers, it is important that the users can understand and visualise the reasons for the decision support predictions. Plausibility can be hard to achieve for complex pathways and models and perceived ’black-box’ functionality often leads to a lack of trust. Here, we describe and evaluate a data-driven framework which moderates some of these issues and demonstrate its applicability to the in-hospital management of community acquired pneumonia, an acute respiratory disease which is a leading cause of in-hospital mortality world-wide. We use the framework to develop and test a clinical decision support tool based on local guideline aligned management of the disease and show how it could be used to effectively prioritise patients using retrospective analysis. Furthermore, we show how this tool can be embedded into a prototype clinical system for disease management by integrating metrics and visualisations for assisting decision makers examining complex patient journeys, risk scores and predictions from embedded machine learning and artificial intelligence models. Our results show the potential of this approach for developing, testing and evaluating workflow based clinical decision support tools which include complex models and embedding them into clinical systems.

## 1 INTRODUCTION

The concept of clinical decision support (CDS) goes back to the 1960s (23) but it was only when digitisation of patient data became a reality in the 1980s that CDS systems became possible. The purpose of CDS is to facilitate optimised and standardised healthcare and as a result modern CDS can vary in complexity from clinical risk score calculators, to medication and patient pathway management and prognostic and outcome modelling. However, options for what is possible within the hospital infrastructure depend on the degree of integration of the electronic patient record (EPR). Where users routinely access disparate or linked systems, CDS provision may become limited or unviable. From a technical and cost standpoint, it can also be difficult to setup and maintain CDS tools. If not embedded at the EPR design stage, CDS tools are often disease/discipline specific, standalone and/or proprietary and not fully interoperable with other information systems (21). Incomplete system integration necessitates duplicate data entry, which not only adds to the healthcare workers’ workload but can introduce transcription errors. The resultant cognitive overload and trigger fatigue effectively prohibits pre-emptive identification and stratification of patients according to clinical risk. This occurs without considering the complexity added by any data processing and lack of system interoperability.

Initially CDS were knowledge-based expert systems driven by a defined set of logical rules to drive the decision making – essentially a series of if/then steps. It is only more recently that non-knowledge based CDS which utilise machine learning (ML) and artificial intelligence (AI) models have been implemented (6). These latter approaches use existing data as a basis for models which identify patterns corresponding to specific predictions -patterns which they can subsequently recognise in any new data presented to them (7). For example, a CDS containing an ML model trained from data containing markers of 30-day mortality for a particular disease, could use this to identify which patients should be prioritised when supplied with new data. However, the growing complexity and scale of AI and ML models used for CDS including access to multi-modal data sources such as images, ‘omics and longitudinal test results into separate systems (as described above) requires prior integration into EPRs to become relevant to clinical management. Additionally, the increased use of non-knowledge based CDS presents ethical concerns. The complexity of these (often proprietary) models makes them appear as ‘black-box’. Understanding these models is required to engender trust in predictions, particularly how such models perform on local data, so that users are encouraged to use the full potential of the system. Perceived lack of transparency, plausibility and trust in accuracy limits generalisability for clinical practitioners who then often prefer to rely on years of knowledge and experience (18).

Here we describe an approach for developing CDS which considers some of the concerns above. This is underpinned by a generic software framework for creating and handling real-time CDS called Embeddable AI and State-based Understandable Logic (EASUL), built based on our experience with management of community acquired pneumonia (CAP) admissions in our local hospital. CAP is an acute lung infection, which is the leading cause of death in UK hospitals, with mortality associated with admissions varying from between 2% and 30% depending on disease severity, comorbidities and age. In addition, it is estimated that annual healthcare costs to the UK National Health Service (NHS) associated with CAP exceed £1 billion. Our previous work showed that prompt interventions are associated with significantly improved outcomes (11) and as a result we wanted to demonstrate how our approach could enable better and more timely use of data and improve prioritisation of patients with more severe disease.

## 2 METHOD

### 2.1 The EASUL framework

The proof-of-concept EASUL framework is based on a Python-based library which enables data-driven CDS plans to be setup and subsequently executed. Figure 1 shows how the EASUL framework can be utilised in different ways to develop CDS plans which contain the data sources, steps, algorithms, states and visualisations representing potential patient journeys. A key advantage of EASUL is that plans can be defined, evaluated and tested using static data sources, before they are adapted into information systems driven through real-time data sources. This is implemented through an optional web API (built using the Django framework), which can serve outputs and results (visualisations and data related to the plan), and a flexible engine supporting use of different broker and client setups to message other systems such as EPRs or databases as required. This approach is generalisable and adaptable to different scenarios and conditions, potentially enabling EASUL to support decision making at any stage of the patient journey.

**Figure 1.**
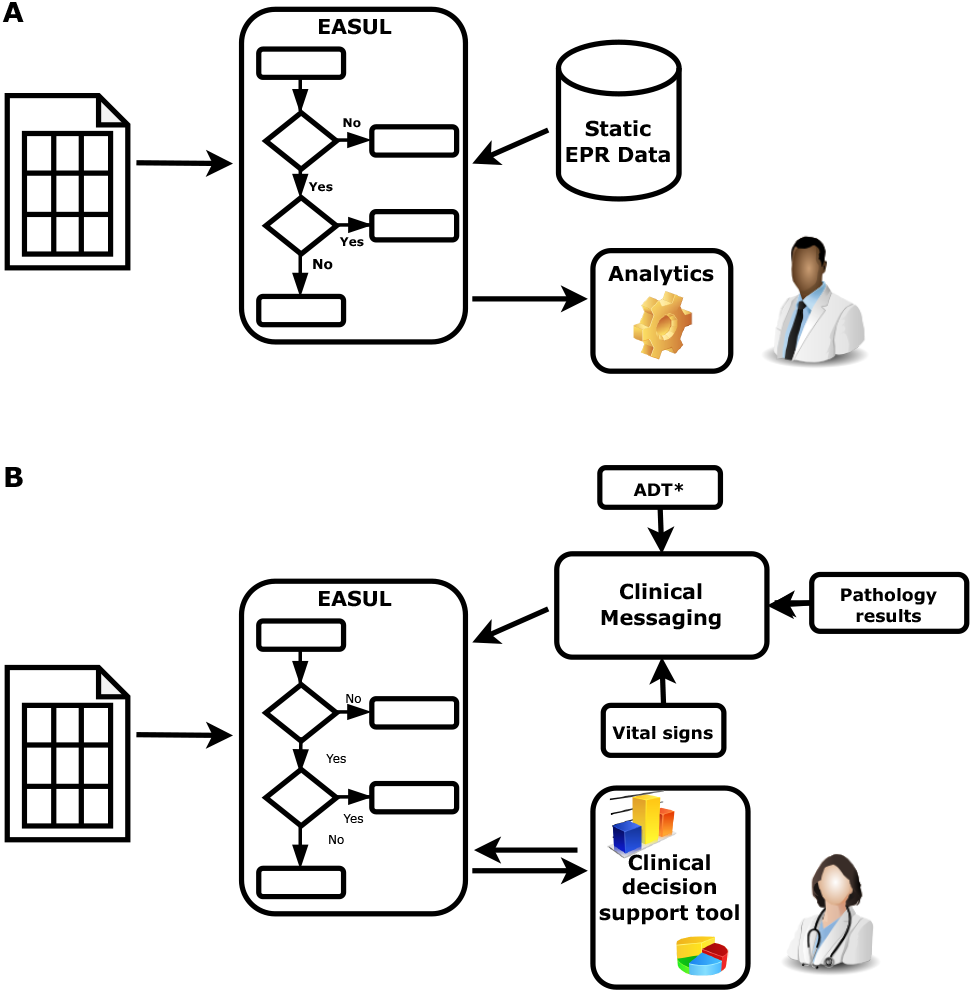
Schematic showing how EASUL was utilised in two different ways. **(A)** For research, quality and service improvement using static data sets and scripting/analytics tools. **(B)** Creation of CDS tools through integration of outputs/results into clinical information systems. *ADT = hospital admissions, discharges and transfers

The main constituent of plans are determinative steps which as shown in Figure 2 support algorithms of different modalities -varying from simple clinical risk scores and logical (if/then) comparisons to advanced ML and AI models – to determine a specific patient journey. To simplify the integration of scores and models we also included data-related concepts in EASUL (schemas, algorithms and data sets) which allow the quick implementation and inclusion of existing models within plans. EASUL also contains tools to serialize algorithms and calculate appropriate metadata (including model performance metrics, model explainers and other values) and specific result contexts which are used to generate appropriate visualisations. To improve performance, the library can pre-calculate and cache metadata and result context to ensure visualisations are provided to the user in a timely fashion. The visualisations are produced using HTML, MermaidJS (5), and matplotlib (13). Information related to models such as area under the curve, sensitivity, specificity and accuracy; and interpretability measures such as explainers are generated using scikitlearn (17). For outputting interpretable elements, the LIME library is used to calculate patient-specific explainers (19) and the results of the explanation are output using plain HTML.

**Figure 2.**
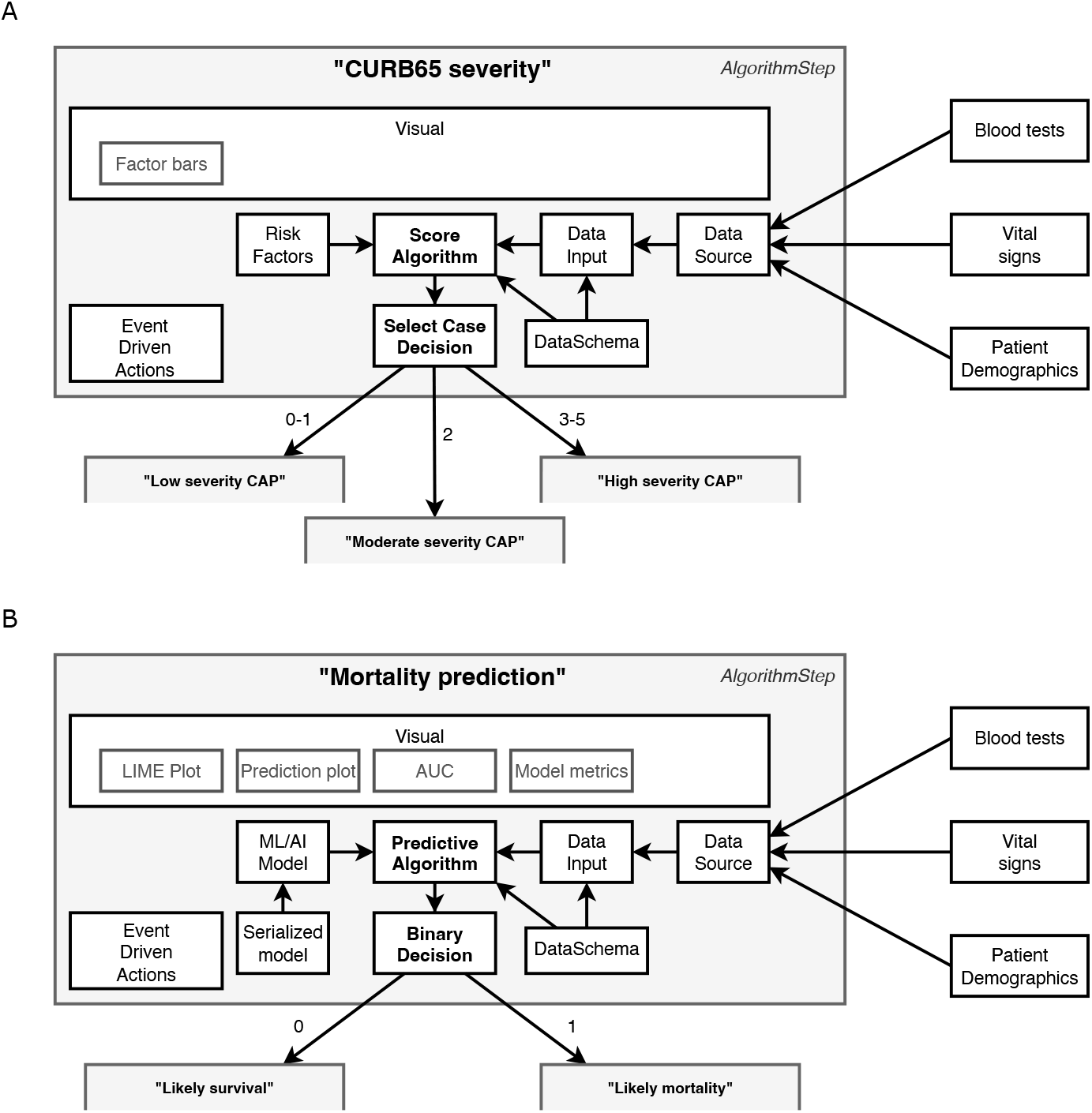
Example components in data-driven algorithm steps. Steps act upon the results of algorithms. This figure shows steps containing **(A)** a ‘Score algorithm’ for CURB65 severity and **(B)** a ‘Predictive algorithm’ for mortality prediction, and while many of the components utilised are the same there are also differences. All algorithms use ‘Data Schemas’ to define the input and output fields which are supported by an ‘Algorithm’ and ‘Data Input’. This allows data to be validated and/or converted before it is used for prediction and prevents a data set being used to make predictions using an algorithm with an incorrect schema. The ‘Data Input’ is collected through collating different sources into a single ‘Data Source’ according to their availability and the system setup. Which step is next in the patient journey is then determined by the results of the ‘Algorithm’ and the ‘Decision’. Decision types are algorithm agnostic, although in **(A)** there are three possible decisions (Low, moderate or high severity), whereas in **(B)** there are two (Likely survival or Likely mortality). There is also flexibility in the event driven actions, which can be set to occur at particular points within the step. For example, one type of action stores a new state value once a decision has been made but before it has been actioned. The main differences between the two steps lie in how the algorithm is defined and, in the visualisations, available. The ‘Score algorithm’ in **(A)** (essentially a risk score) is built from ‘Risk factor’ expressions, whereas the ‘Predictive algorithm’ in **(B)** comprises a previously trained and serialized machine learning/AI model – both powered by ‘Data Inputs’.

The current version of EASUL is available for download, along with documentation at https://github.com/rcfgroup/easul.

### 2.2 Data sources

To develop and evaluate the CAP management plan, individual hospital episodes from adults (≥ 16 years old) admitted between April 2022 and June 2022 were extracted from the hospital data warehouse as delimited text files. Alongside this, records of patients reviewed by the SPIN team were extracted from a locally run service database. The hospital data included demographics (patient ID, admission ID, dates and times of admission/discharge and the nature of the discharge) as well as International Statistical Classification of Diseases and Related Health Problems version 10 coded primary and secondary diagnoses, blood test results, oxygen records and drug records. The SPIN data included patient ID, dates/times of admission and CURB65 severity calculated by the SPIN team. Data files were de-identified prior to loading into a SQLite database for subsequent evaluation.

### 2.3 Decision model development and evaluation

A CAP management plan was defined in Python containing the appropriate steps and associated properties. The plan also included data transformations/processes so that EASUL would convert and process data in non-standard formats on-the-fly. This included date/time parsing and CAP diagnosis based on an algorithm utilised previously (11) but refactored into an EASUL compatible process. For this development and evaluation, the SQLite database described above was established as the main data source in this plan.

To evaluate the utility of the CAP management plan two data sets were created. The first data set comprised data derived from executing the EASUL plan across all the admissions in the SQLite database to determine diagnoses, patient journeys, derived states and algorithm results. The second data set was created by extracting the SPIN review data from the SQLite database and linking it to the coded admission records through an anonymised patient ID and date/time matching algorithm, with admissions not reviewed by the service identified and filtered out. This resulted in two sets of delimited text files representing the CAP admissions: one containing the EASUL-related data for all admissions, including those determined to have CAP based on the above CAP diagnosis algorithm and the other admissions reviewed by the SPIN service.

The two data sets were merged and then appropriate variables compared using Python and the pandas library. The primary comparison undertaken in our evaluation was between the level of severity recorded by the SPIN team and that determined by EASUL. As defined previously, in the UK CAP disease severity is classified using CURB65 as low (CURB 0-1), moderate (CURB 2) or high (CURB 3-5) (1). While EASUL automatically calculated CURB65 and the derived severity level for each hour of each CAP admission, the SPIN data only contained a single record of a manually calculated CURB65 score. Therefore, we used this score to categorise severity and then compared it to the highest severity level determined by EASUL during an admission. The frequency of each severity pair (e.g. low-medium, high-high) was tallied. The other evaluation we undertook was to examine drug concordance with severity. Individual drug records for each admission were summarised according to a binary flag marking the presence/absence of specific drugs used to treat CAP and compliance was then determined by looking at the presence of specific drugs corresponding to severe (high) or non-severe (low or medium) disease, and comparing this to the EASUL severity level.

### 2.4 Proof-of-concept CDS system

Our CDS implementation utilised the Headfake library (10) as the basis of a bespoke in-silico “simulated admissions” tool. This output near real-time data matching the record/field formats obtained from the hospital data warehouse and pertaining to admissions/discharges and chest X-ray reports, pathology tests and vital signs -with the records encoded as simple JSON structures and being sent as messages at times during each ‘admission’. The EASUL plan was also altered to accept near real-time data from the broker and to embed an ML classifier from an ongoing project which predicts whether patients had died in-hospital or within 30 days of discharge from CAP (15). This classifier, based on tree gradient boosting and built using XGBoost (8) was wrapped as an EASUL algorithm along with a data schema and encoder/decoder designed to process routine hospital data into the correct format of data for input into the model. Additionally, visualisations were also defined representing this algorithm for both the overall model (e.g. performance metrics) and the row (e.g. breakdown of probabilities and other measures of interpretability). Due to the headless nature of EASUL, to provide an appropriate user interface for the CAP management system, we used a local bespoke clinical data collection platform and built a CAP data collection/management module and custom extensions to handle near real-time data from the EASUL broker.

## 3 RESULTS

### 3.1 Creating and evaluating EASUL for CAP management

To demonstrate the potential utility of EASUL for CAP management CDS we worked with local physicians and members of the SPIN team to identify the key steps and decision points in the current person-driven service process (11) and used EASUL to set this up as a data-driven workflow plan for CAP management.

The utility of this plan was assessed through a local quality improvement (QI) project. This involved comparing the results obtained from feeding retrospective admissions data into EASUL, with the actual events and information recorded by the service team. The admissions data set consisted of 52,471 adult admissions, with 630 diagnosed retrospectively with CAP at admission.

Figure 3 shows that 283 (44.9%) of the CAP admissions were reviewed by the SPIN team. Of these it was possible to compare the severity determined by EASUL and the SPIN team for 227 (80.2%). Only 112 (49.4%) had the same severity determined by both EASUL and the SPIN team, which meant that over half did not match, with 57 (25.1%) of reviewed admissions identified by EASUL as high severity but recorded only as low or moderate severity by the SPIN team. Conversely, 5 (2.2%) of the admissions were classified as moderate severity by the SPIN team, but deemed only low severity by EASUL. Crucially for the approach, no patients were classified as high severity by the SPIN team and as low severity by EASUL. To confirm the comparison was correct, the data from a proportion of the patients who were classified differently by EASUL and the SPIN team was examined manually. Based on this it appeared that some of the patients had indeed been incorrectly scored by the SPIN team. There are several reasons why this could be the case including data missing / missed during the admission; results arriving or being recorded later due to clinical pressures; or patients being discharged prior to review. Additionally, exactly how and when CURB65 is applied is not always systematic and can be based on clinical judgement. Finally, it is also important to note that 347 (55.1%) of the CAP admissions were not reviewed by the SPIN team, with 244 (70.3%) of these deemed to have had moderate or severe disease.

**Figure 3.**
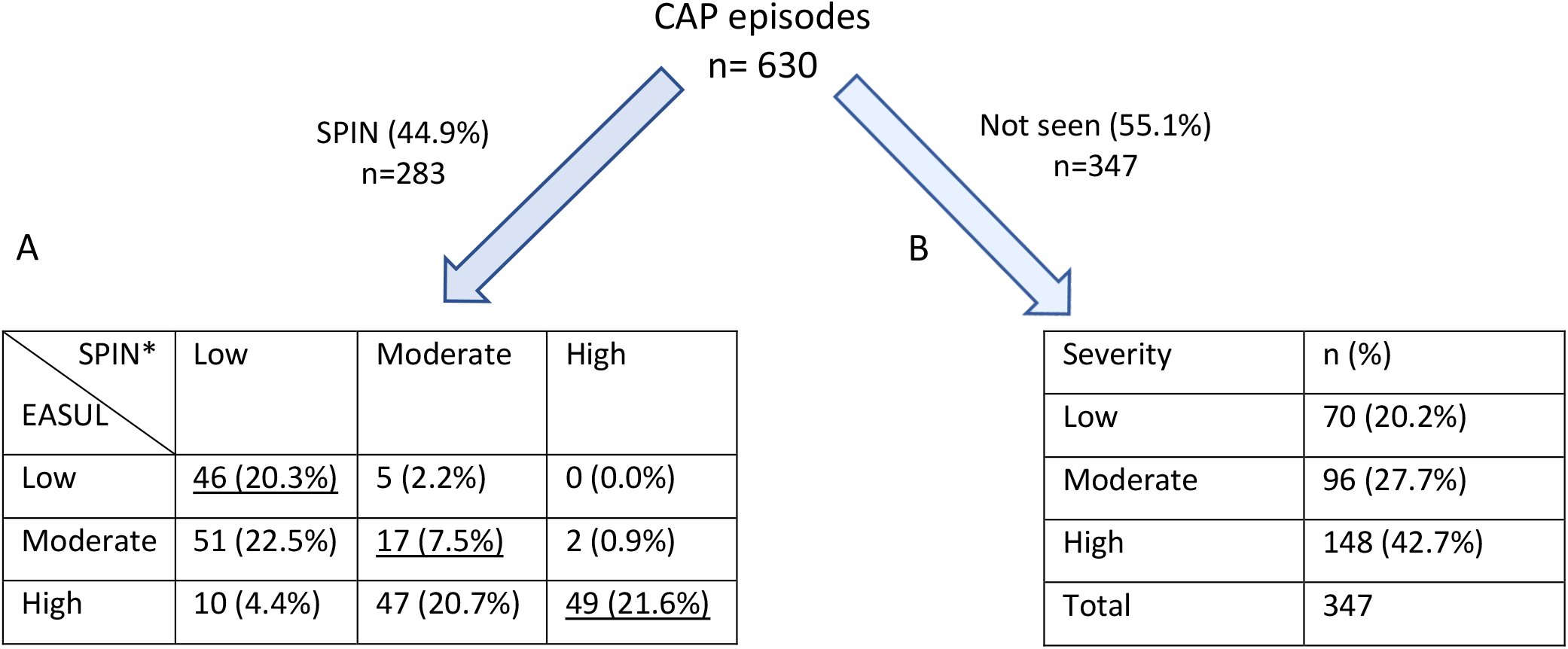
Number of admissions dichotomised into those seen / not seen by SPIN showing calculated severity levels. **(A)** Cross-comparison of severity automatically calculated by EASUL with those manually calculated by the SPIN team. **(B)** Severity automatically calculated by EASUL. Underlined values match in severity. *CURB65 was only recorded for 227 admissions seen by SPIN. Severity level is based on: Low = CURB65 0-1; Moderate = CURB65 2; High = CURB65 3-5.

Our secondary evaluation looked at drug concordance with severity (low-moderate vs high) in 591 (93.8%) of the CAP episodes where drug data was available. This indicated that 503 (85.1%) were prescribed drugs concordant with high severity CAP, despite patients having only low or moderate severity disease; while only 88 (14.9%) had the correct prescriptions according to concordance.

### 3.2 Embedding decision support into a clinical tool

In addition to showing how it could improve prioritisation of CAP admissions, we also wanted to demonstrate how our framework could be used to develop embeddable CDS tools. Figure 4 shows a proof-of-concept architecture for achieving this. A key component of this is a ‘simulated admissions’ tool which imitates patient flow by generating near real-time synthetic data. This pushes appropriate simulated data to the Broker at corresponding time points which drives the Handler service, utilising the HTTP Client to progress the patient journey accordingly. When no data is available at a particular step in the plan, or the journey reaches completion (e.g. the patient is discharged or dies) the Handler pauses and either awaits further data or finalises the journey. During the process, EASUL outcome data is sent back to the broker to feed the Receiver service in the Clinical System. This leads to the creation of new or updated clinical records according to where a patient is in their journey. The Clinical Frontend can also access and display data and visualisations from the Client related to specific journeys. The results of this are shown in the screenshots in Figure 5 which demonstrates a prototype Clinical Frontend based around a prioritisation dashboard with embedded visual elements and results from the EASUL plan for CAP management.

**Figure 4.**
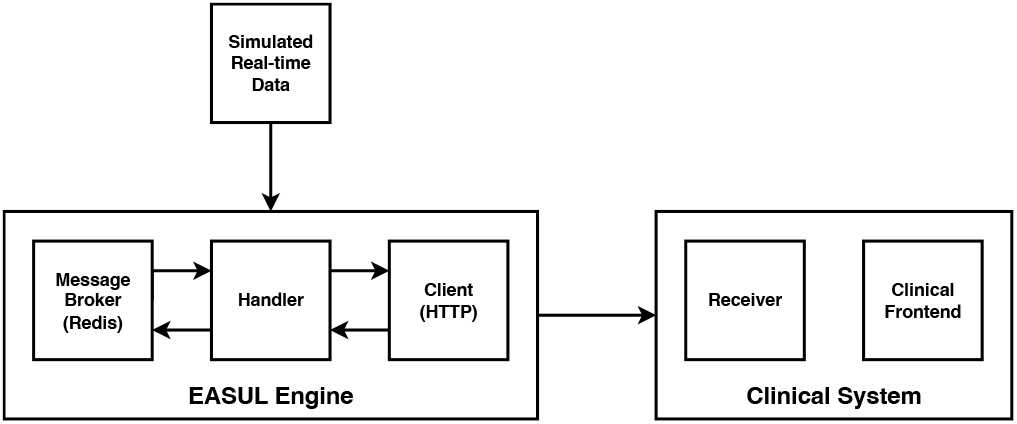
High-level architecture of the proof-of-concept CDS system incorporating simulated real-time data.

**Figure 5.**
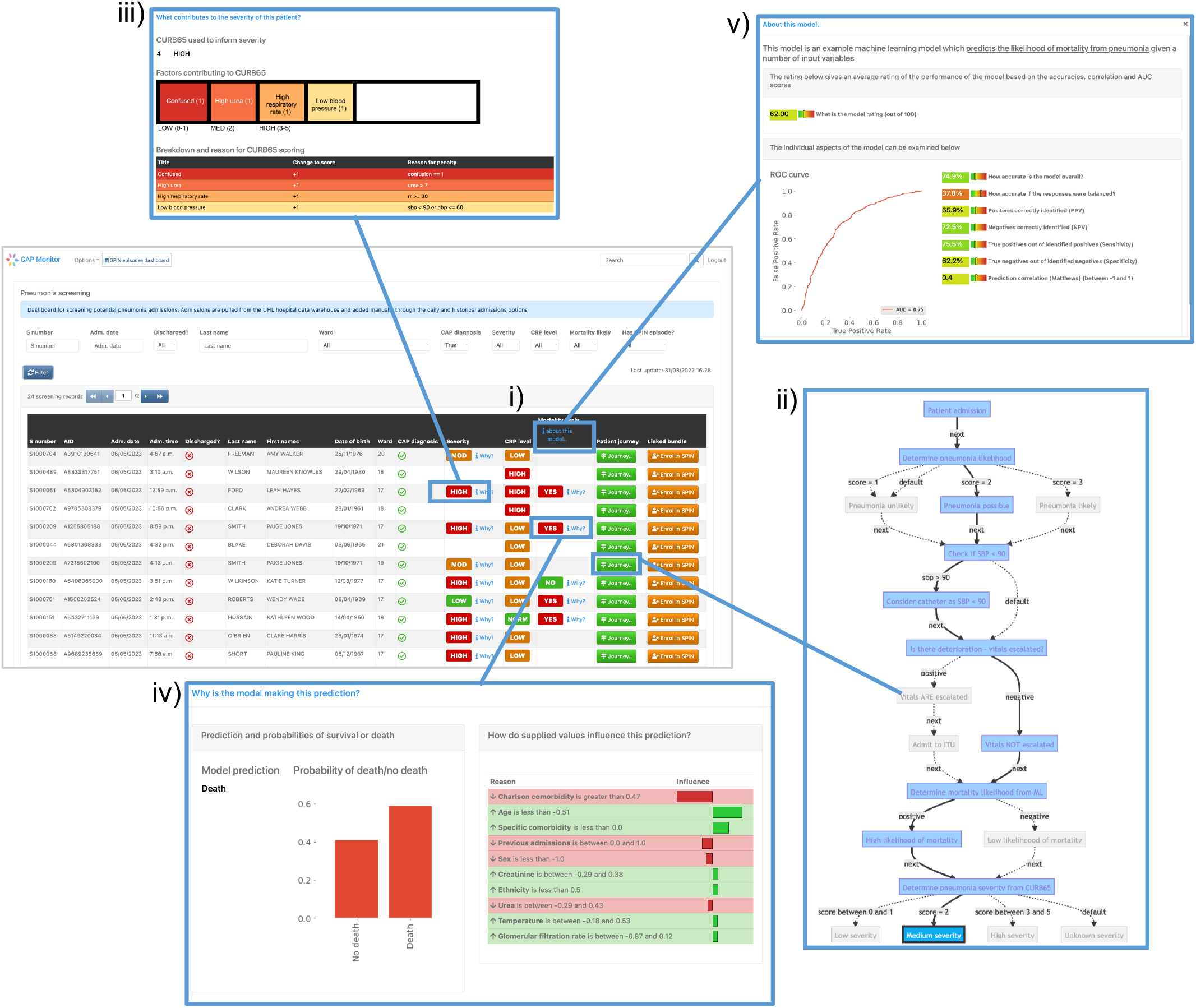
Screenshots of the prototype clinical frontend showing the features provided by EASUL. Admissions are added automatically to the system once a patient is identified as having CAP according to presence of specific clinical codes (see Methods). The resulting dashboard contains links to several visualisation options to provide support to the user: i) results from CURB65 (severity) and CRP level stratification, along with an ML model to predict likelihood of mortality; ii) visualisation of the journey so far for the selected admission, showing key decision points driven by the data. This can be customised to include data sources and/or only show the direct route; iii) breakdown of the CURB65 severity score into individual risk factors; iv) visualisations to further explain the model predictions with a bar plot showing the prediction probabilities and interpretable outputs such as LIME plots showing the variables and values which influence the prediction; v) model overview which provides key model performance indicators including area under the curve, accuracy, predictive power, sensitivity and specificity.

As part of our prototype, we also embedded an ML model which predicted pneumonia outcome. This gave us the opportunity to implement visualisations to improve on interpretation of specific results output by the model (e.g. LIME-based plots) and present the reasons for predictions, along with metrics showing model performance, such as ROC (including area under the curve), accuracy, recall and precision.

## 4 DISCUSSION

We demonstrate here how our approach can effectively be used in operations, research and decision support tool development. Through our QI project we were able to show its utility for prioritising and triaging patients who may not otherwise receive the care required in a timely fashion. As we previously reported (11), patients not reviewed by the SPIN service have notably poorer outcomes, most likely because they are prioritised less effectively. Early identification of the *>* 200 high priority CAP admissions not reviewed by the service suggests that better use of data through EASUL-type approaches would improve this situation, particularly if used as a triage to screen and exclude lower priority patients. We believe that tools like the prototype CDS we demonstrate would have significant utility in clinical practice and envisage many situations where they could benefit patient outcome through automation and simplification of straightforward data-driven processes in hospital patient management to identify earlier treatment opportunities.

Our interest in pneumonia management was driven by a need for equitable access to evidence based interventions across a large multi-site healthcare organisation. Local QI priorities facilitated access to appropriately granular severity data. The very experienced and database enabled SPIN team played a key role in promoting the quality of this dataset as relevant to outcome prediction. Additionally, the team’s collection of secondary qualitative data facilitated this comparative quality improvement project. Cognisant of more advanced pneumonia-focussed decision support tools in active use (9) our main objective here was to develop and evaluate a generic tool which could be utilised in similar acute hospital trusts, and we are not comparing our CAP solution with more fully featured single-purpose CDS tools. While we considered using an existing freely available dataset to demonstrate our concept, we were keen to apply the tool to a local real-life acute respiratory emergency care scenario and data set rather than a hypothetical one, thereby accepting some additional complexities and limitations.

Existing generic approaches for implementing web-based interfaces for statistical models and processes (e.g. RShiny) are not focussed on data and system integration and generally entail entering input data into a user interface. While our project does have some similarities with other projects, there are key differences. Leiner et. al describe a “vendor-neutral” AI infrastructure although their solution is specifically targeted towards imaging (14); while the creation and deployment of AI-based models as CDS is a key focus of the KETOS platform (12). There have also been other projects which focus on knowledge-based CDS run as microservices (12) and semi-automation of multi-modal AI model development (20). However, unlike EASUL these do not appear to include workflow-driven approaches and customisable visualisations or support different modalities of source data.

Our proof-of-concept clinical system allowed us to demonstrate how algorithms of varying complexities could be embedded into an information system in a re-usable way. Using EASUL we were able to include both simple risk scores and a pre-existing ML model in a real-time data-driven workflow. This is significant since ML models can be effective, but are not always simple to integrate into such workflows. Additionally, we included both model-based and patient-based visual elements to help with interpretability, accountability and understandability. There is a growing interest in this field and we agree with others (21, 24) that transparency and accountability around both the strengths and weaknesses of ML and AI is crucial for gaining the trust of clinicians when using CDS based on these models, particularly given the vast increase in the number of published predictive models (16) and continuing advances in the field. However, we acknowledge that this must be done carefully to ensure that increased transparency does not compromise the security of models/algorithms and lead to potential increases in bias (25).

The flexible nature of our approach means it can be extended to support different data modalities (e.g. ‘omics data, imaging); adaptive workflows including multi-tiered/decision models and potentially mobile applications. Additionally, it could also be used to support patient directed healthcare actions, such as remote monitoring. From a technical perspective our use of generic plans makes it straightforward to modify the approach and allows parts of plans to be re-used, while other aspects can be customised as needed in the same project (e.g. for separate evaluation and implementation). The framework also includes support for storage and re-use of previously trained models, with input data validation based on a data schema and different extraction-transform processes, including the ability to collate different data sources together and to have custom extraction-transform processes applied at the point records are received. While this has some similarities to approaches such as FIDDLE (22), our method is data source and format agnostic. However, we recognise its current lack of support for standards such as Health Level Seven and Fast Healthcare Interoperability Resources (4) is a limitation and are keen to add this due to a growing interest in the UK NHS and elsewhere (3, 2).

The work presented here is preliminary research to establish the potential of our approach and we fully acknowledge that further development would be required to improve security, resilience, and scalability. Despite this, we believe that EASUL and similar approaches are important steps in making better use of health data from multiple sources and would help to strengthen trust and accountability in complex ML and AI model enabled CDS. Further research and development in this area is required to evaluate and utilise the available data and models being developed in clinical practice and produce digital health tools which are fit for use in clinical settings.

## AUTHOR CONTRIBUTIONS

RF, GW and PH contributed to conception and design of the study. DL developed the machine learning model used in the prototype CDS. JS and LS led and undertook data acquisition as part of the SPIN team. DL and MS provided intellectual contributions. RF wrote the first draft of the manuscript. All authors contributed to manuscript revision, read, and approved the submitted version.

## ACKNOWLEDGMENTS

The authors would like to thank the SPIN team, past and present for their assistance during the project and Mei-Mei Cheung and other members of the Research and Innovation team at the University Hospitals of Leicester NHS Trust for providing data exports for the evaluation.

## DATA AVAILABILITY STATEMENT

The EASUL framework is licensed under the GNU Lesser General Public Licence v3.0 (LGPL-3.0) and is available at https://github.com/rcfgroup/easul. The evaluation presented in the manuscript was part of a quality improvement exercise, therefore we do not have ethical approval to share the data used to produce these results.

## ETHICS STATEMENT

The conduct and reporting of the evaluation in this project were discussed with the local research governance committee and in line with the Policy Framework for Health and Social Care 2017 a formal ethical review was not required.

